# Identifying weather patterns affecting household date palm sap consumption in Bangladesh, 2013-2016

**DOI:** 10.1101/2024.05.06.24306951

**Authors:** Jules Jackson, Ireen Sultana Shanta, Clifton McKee, Stephen P Luby, Najmul Haider, Yushuf Sharker, Raina Plowright, Peter Hudson, Emily Gurley

## Abstract

Nipah virus spillovers via consumption of date palm sap in Bangladesh vary substantially between years and have been associated with lower winter temperatures and precipitation. However, the mechanisms driving the interannual variation and the influence of weather remain unexplained. Here we investigated the hypothesis that weather patterns change human sap consumption and explain interannual variation in observed spillovers. We analyzed responses from a nationally representative survey conducted among 10,000 households in Bangladesh in 2013-2016 on household date palm sap consumption and weather data for each division of Bangladesh, using logistic regression to examine whether sap consumption is associated with weather variability. We found significant associations of lower minimum temperatures and precipitation with increased household sap consumption during the sap harvesting season. This relationship was largely similar within all months and divisions, and strong associations of temperature (**χ**^2^ (1, n =5,027) = 7.74, *p* < 0.01) and, independently, precipitation (**χ**^2^ = 8.00, *p* < 0.01) remained strong after accounting for month, location, and annual sap season. Interannual variation in date palm sap consumption in Bangladesh is likely best explained by temperature and precipitation patterns, where colder, drier winter days pose a higher risk for Nipah virus spillover. The synthesis of approaches to determine the driving forces of seasonality in our study may be a valuable method for investigating seasonality in other zoonotic pathogens.

## Introduction

Date palm sap consumption is a longstanding practice in Bangladesh and nearby regions in South Asia, where date palm trees have been cultivated for centuries for their sap [1]. The practice was linked to Nipah virus spillovers after in-depth interviews and case-control analysis during an outbreak in 2005 [2] and drinking sap has been a consistent risk factor for human Nipah virus infection through years of case-control risk factor analyses [3, 4]. Nipah virus case fatality is approximately 70% in South Asia and person-to-person transmission commonly occurs [5], posing a risk for highly fatal regional outbreaks, or possibly pandemics. Bangladesh has reported more spillover events than any other country since its first reported human cases in 2001 [6]. By better understanding the factors that contribute to Nipah spillovers we can better identify key targets for intervention that can prevent future cases of Nipah virus spillovers, and subsequent outbreaks.

Though date palm sap is tapped year-round for various purposes, the sap is only consumed raw in the winter months between November and April when the temperatures are sufficiently low to keep the sap fresh, herein referred to as the sap season. Bats are thought to introduce the virus into the sap through shedding from bodily fluids (saliva and urine) while consuming the sap during the tapping process [7]. However, current understanding of the nuances of sap consumption in humans remains limited.

Nipah virus spillovers in Bangladesh have occurred more frequently during years with colder winter temperatures [8]; furthermore, investigation has shown weekly minimum temperatures to be a strong predictor of spillover [9]. Several hypotheses have been suggested to xplain this correlation. One hypothesis is that the virus may survive longer in colder and drier conditions. However, examinations of Nipah virus survival in fruit juices with a comparable pH to date palm sap showed a survival time of several days at temperatures warmer than those observed in Bangladesh during the November–April sap harvesting season [10].

A second hypothesis is that colder temperatures could be affecting the behavior of the reservoir, *Pteropus medius*. The stress of colder weather may impose increased energy demands on bats, and the availability of the fruits that compose the majority of their diet is reduced during the winter months. These conditions may lead bats to turn more frequently to a reliable food source with a high sugar content such as date palm sap when it is available, especially so during the coldest days of the sap season [7].However, though bats did access sap more frequently during the cooler winter and springtime months, the frequency of *P. medius* visits to date palm trees in winter was not significantly related to temperature on a daily level in a three-year study [11].

A third hypothesis is that human date palm sap consumption changes based on temperature. A recent study identified significant differences in the frequency of household sap consumption from year to year. One potential explanation is that humans drink more sap during the coldest days of the winter when sap is more abundant and they report that the sap is sweetest [12, 13] (Fig 1). More recently, lower precipitation in the month preceding the spillover event was also observed to be a predictor of spillover risk, along with El Niño Southern Oscillation cycles with a lag of three months [14]. One speculative explanation for this is that rain during the tapping process may dilute the sap, making it less sweet and less appealing to buyers. Of all the proposed hypotheses for the weather-spillover relationship, these have received the least attention.

**Fig 1.**
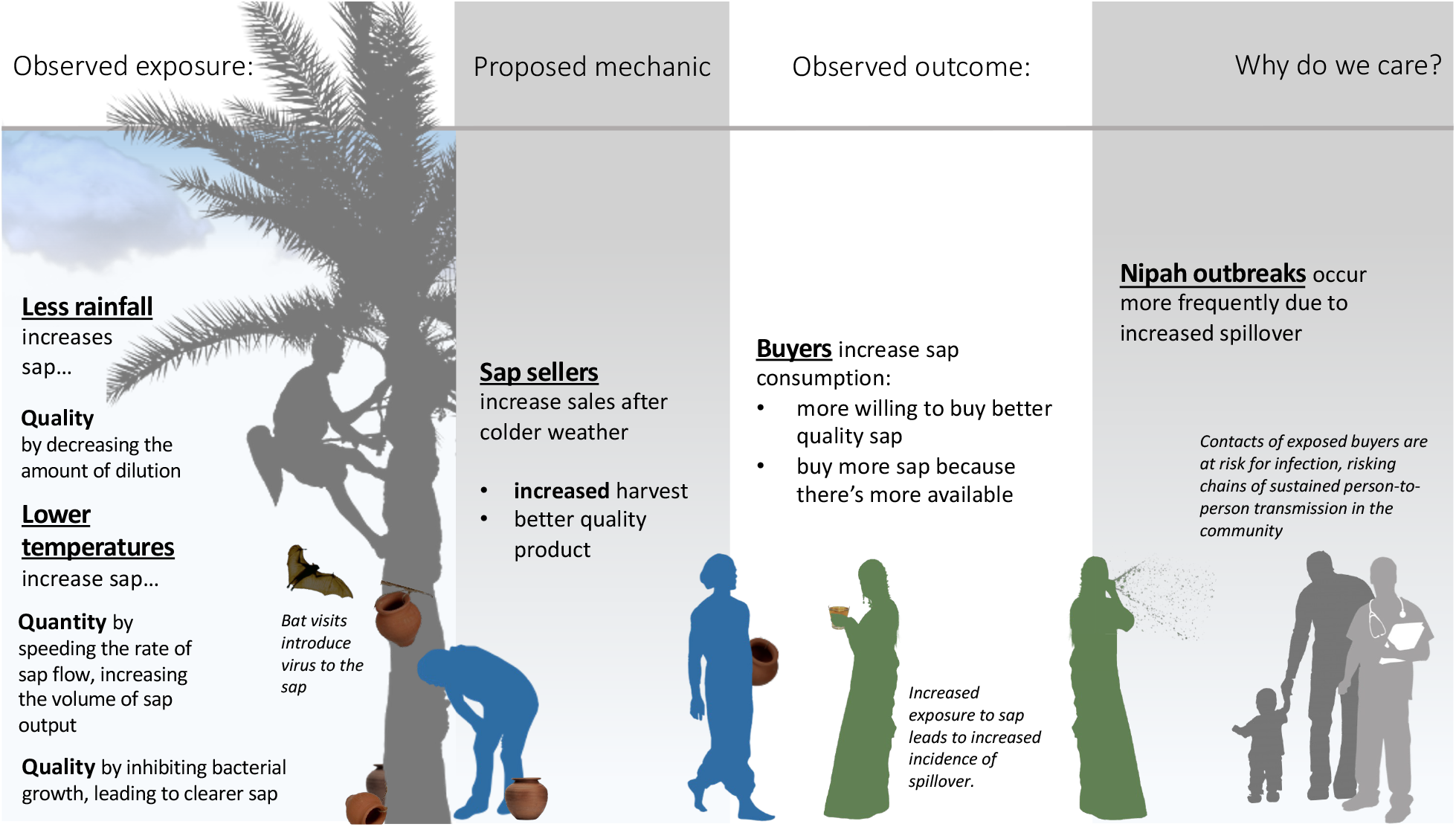
Proposed Mechanics of Weather-Spillover Relationship. Proposed relationships between winter temperatures, precipitation, seasonal sap production, sap consumption, and Nipah virus spillover risk.

In this study, we aim to examine associations between weather patterns in Bangladesh and household date palm sap consumption in Bangladesh 2013-2016.

## Methods

### Sap consumption

To investigate changes in date palm sap consumption over time, we used data from a nationally representative household survey of adults in Bangladesh performed between September 2013 and August 2016. Additional details on study design and sampling strategy can be found in Shanta et al., 2023 [15]. The study was conducted in 10,002 households in 1,001 communities representing both urban and rural populations across the 7 administrative divisions of Bangladesh. Communities were randomized within 16 strata of population size in all administrative divisions each season of the study period (winter: November–February, summer: March–June, and rainy: July–October) to ensure comparability in terms of number of communities enrolled between each season and location for the duration of the study. Households were identified by a modified random walk method, using the main entrance of the homestead with the most recent wedding in rural communities, and ward entrance as random starting points for urban communities. In rural areas, the house to the left of the main homestead was enrolled first, then every third household was approached in that direction for enrollment until 10 households had been enrolled. In the case that the household declined study participation, each successive household was approached. In urban areas, surveyors chose dwellings by randomly selecting from envelopes containing cardinal directions in each sampled ward. Teams then followed the direction until reaching an administrative boundary of the ward, selecting the first dwelling at the boundary, then approaching every fifth dwelling by proximity for enrollment. Participants were selected from adults within households for their willingness to discuss select household activities that occurred in the past month relative to the date of the survey, including the harvesting and drinking of date palm sap by household members. Participants without a recorded response and those who responded “Don‘t know” were excluded from analysis.

### Weather data

To investigate the association of weather and date palm sap consumption, weather data were retrieved from 10 weather stations in Bangladesh included in the National Centers for Environmental Information’s (NCEI) Past Weather database (https://www.ncei.noaa.gov/past-weather/), managed by the National Oceanic and Atmospheric Association (NOAA) (S1 Fig.) The administrative division of each weather station was identified from geographic coordinates provided by the NCEI’s Historical Observing Metadata Repository (HOMR) location information. Further weather data were acquired from the Bangladesh Meteorological Department. Weather data included daily summaries of average, minimum, and maximum temperatures, cumulative precipitation, dew point temperature, surface pressure, and visibility from July 2013 through December of 2016. When multiple stations were included within a single division, daily summary values were averaged across the stations and the resulting mean daily value within that division was used. Daily values for relative humidity were calculated using daily values for dew point and temperature from each station across Bangladesh.

### Analysis

Monthly averages of daily values of average temperature, minimum temperature, cumulative precipitation, relative humidity, and visibility were calculated over the entire study period. Because survey responses were obtained continuously throughout months, monthly averages were calculated as both categorical (i.e., the average of daily values in the division over the month attributed to all responses in the division that month) and as a moving average. The moving average was determined using the daily observations within the division over the 30 days preceding the response to better cover the time period directly relevant to the survey respondents’ household sap consumption (“In the past month…”). When there were insufficient observations to calculate a moving average for that day and location, values were imputed with the last observed value carried forward within the division. For each response, both the categorical monthly averages and 30-day moving averages of weather variables were examined.

We first investigated the association between sap consumption and weather during the sap season by summarizing consumption across locations, months, and sap seasons. Monthly sap consumption was treated categorically outside of regression analysis. All data analysis and visualization was performed using R (v4.3.1) [16].

Date palm sap consumption is highly seasonal and limited to the months of November-April; we therefore limited our further statistical analysis to responses from those months to avoid effect size overestimates of the association between date palm sap consumption and weather due to the seasonality of consumption. For our analysis, we focused on the association between household sap consumption and minimum temperature rather than average temperature for purposes of comparability with previous analyses by McKee et al., 2021 [8] and Cortes et al., 2018 [9], as well as to better reflect the nighttime conditions during which sap is allowed to flow before harvest in the morning. For each response, we explored both categorical monthly averages and moving 30-day averages of weather variables.

We used likelihood ratio **χ**^2^ tests to compare relative fit between null and extended models of sap consumption to determine how weather contributed to model fit past the expected effects of location and month. As significant relationships between division, month, and household sap consumption were previously identified [3], those two variables were included in our null model. We also included sap season in the null model to account for the possibility of year-to-year differences in sap consumption due to previous interventions on sap consumption. Extended models included categorical and moving averages of minimum temperature, cumulative precipitation, visibility, and relative humidity to determine whether interannual variance in weather conditions contributed to explaining patterns of seasonal sap consumption beyond the expected geographic and monthly differences.

To further explore the possibility that variation in sap consumption may be due to seasonal differences such as changing attitudes towards sap consumption, we used receiver operating characteristic (ROC) curves of weather and season to determine their ability to classify predicted probabilities of date palm sap consumption. We performed model selection to determine the combination of categorical and weather variables that resulted in the best fit using Akaike’s information criterion, corrected for the small sample size of the outcome (AICc).

## Results

Seven percent (361/5024) of respondents reported household sap consumption in the past month during the sap season (November-April). Data on sap consumption during the sap season was missing for 3 households.

Reported consumption was highest during the 2013-2014 season, with 12% (193/1675) of respondents reporting consumption (Table 1). Peak sap consumption occurred in January and February in each sap season of the study (S2 Fig), with 15% (124/857) and 15% (102/698) overall.

**Table 1.** Geographic and temporal characteristics of date palm sap consumption during the sap consumption season (Nov.-Apr.) in 10,002 households in 1,001 communities across Bangladesh, 2013-2016.

| Response characteristic |  | Households reporting date palm sap consumption in the past month |  |
| --- | --- | --- | --- |
|  |  | % | (n/N) |
| **Season** | 2013-2014 | 12 | (193/1675) |
|  | 2014-2015 | 6 | (91/1670) |
|  | 2015-2016 | 5 | (77/1679) |
| **Month** | November | 1 | (3/583) |
|  | December | 5 | (50/1075) |
|  | January | 15 | (124/857) |
|  | February | 15 | (102/698) |
|  | March | 7 | (62/881) |
|  | April | 2 | (20/930) |
| **Location** | Barisal | 5 | (38/720) |
|  | Chittagong | 4 | (30/720) |
|  | Dhaka | 5 | (39/718) |
|  | Khulna | 18 | (126/717) |
|  | Rajshahi | 13 | (96/720) |
|  | Rangpur | 3 | (21/719) |
|  | Sylhet | 2 | (11/710) |
| **Total:** |  | **7** | **(361/5024)** |

There was an inverse relationship between sap consumption and minimum temperature during each sap season, though the strength of the relationship between consumption and minimum temperature varied by season, from weakest during the 2013-2014 season (slope = - 6.97) to strongest during the 2015-2016 season (slope = -24.08) (Fig 2A). Though consumption is lowest at the margins of the sap season (November and April), there were relatively high levels of consumption during the November, March, and April of the 2013-2014 season as compared to the 2014 -2015 and 2015-2016 sap seasons (S2 Fig).

**Fig 2.**
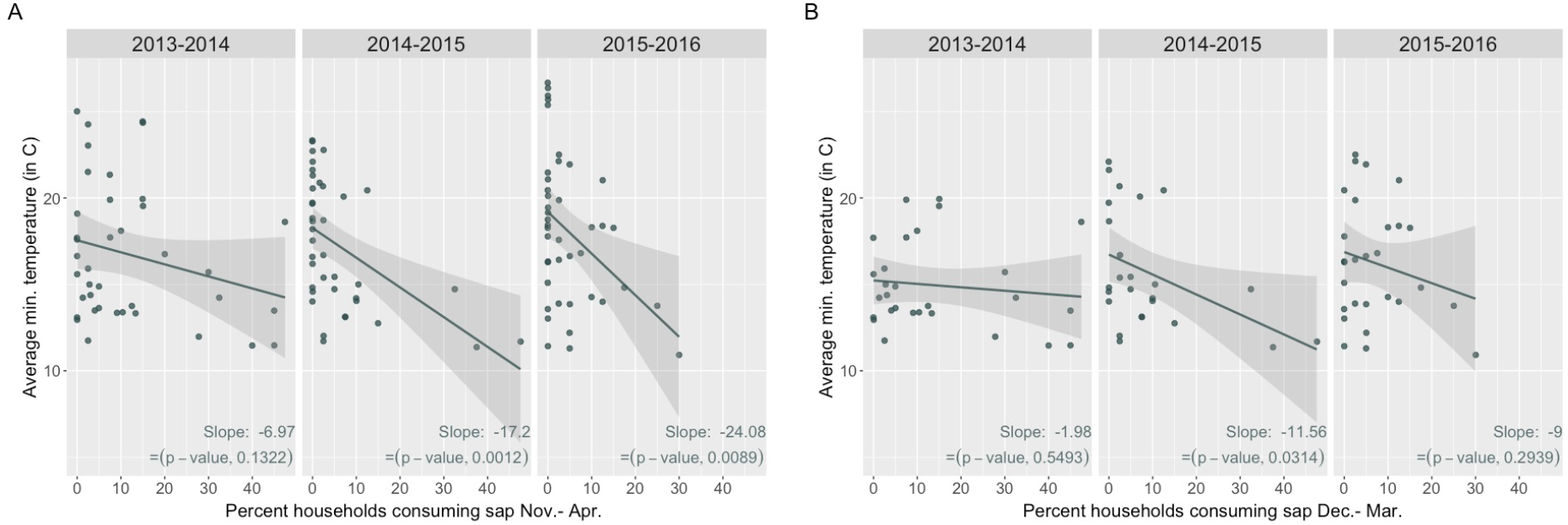
Sap consumption and minimum temperature by season. (A) Monthly percentages of household sap consumption and monthly average of daily minimum temperatures by location over sap seasons, Nov.-Apr. (B) Monthly percentages of household sap consumption and monthly average of daily minimum temperatures by location over a subset of months within the sap season, Dec.-Mar.

To account for the possibility that the relationship between monthly percentages of household sap consumption and minimum temperatures by location was impacted by the high late-fall and spring temperatures in the November and April of the sap season, we further examined the relationship after limiting the analysis to December-March (Fig 2B).

Sap consumption remained negatively correlated with average minimum temperatures during each sap season of the study period (slopes -1.98, -11.56, and -8.99, respectively), even after restricting to the coldest months with the most sap consumption.

The relationship between average monthly minimum temperatures and monthly percentages of household sap consumption was largely similar across all administrative divisions (Fig 3), exhibiting a consistent inverse relationship between sap consumption and monthly averages of daily minimum temperatures. However, districts with low overall average sap consumption, particularly Rangpur (3%), Chittagong (4%), and Sylhet (2%), did not demonstrate this association to the extent that districts with greater sap consumption did.

**Fig 3.**
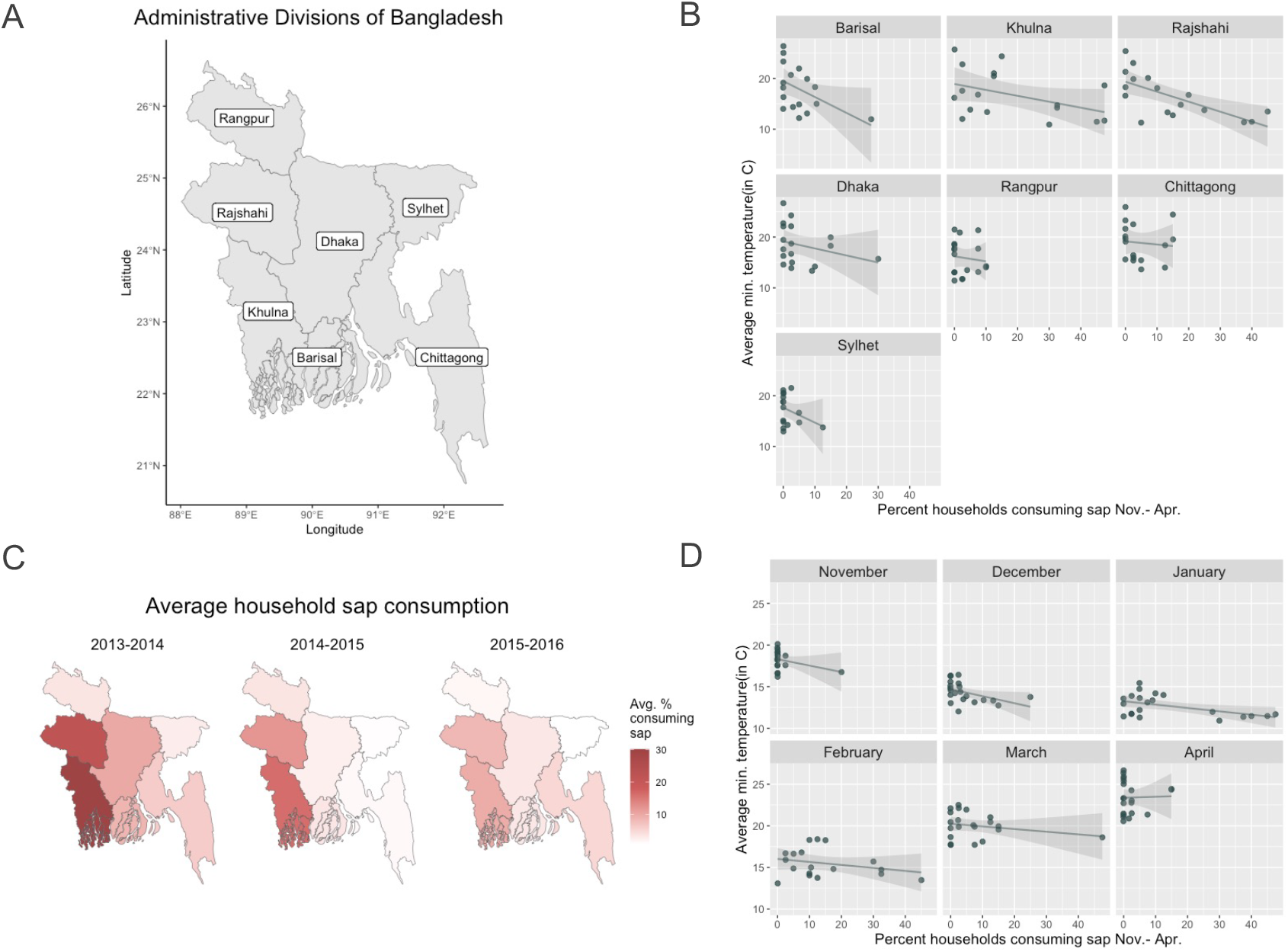
Sap consumption and minimum temperature by month and location. (A) Map of administrative divisions of Bangladesh, (B) Monthly percentages of household sap consumption and monthly average of daily minimum temperatures by location over all sap seasons of the study period, Nov.-Apr. 2013-2016. (C) Average percentages of household sap consumption across sap seasons in each division and (D) Average percentages of household sap consumption and minimum temperatures in each division by month across all sap seasons.

The peak time for sap consumption varied by location over the study period (Fig 3C). The peak occurred in January for Barisal (13%), Chittagong (8%), and Khulna (41%), while the peak of the season was in February for Dhaka (16%), Rajshahi (31%), Rangpur (7%), and Sylhet (9%). We found an inverse relationship between average monthly temperature and monthly proportion of sap consumption nearly each month of the sap season with the exclusion of April (Fig 3D).

We observed greater household sap consumption in the 2013-2014 sap season (Fig 4A), as well as colder monthly averages of daily minimum temperatures (Fig 4B) and lower monthly averages of daily cumulative precipitation (Fig 4C) in the 2013-2014 sap season. Further, minimum temperatures and sap consumption were related by season and location, with not only colder years in general but colder winters in a division being related with greater sap consumption in that division. No association was identified between household sap consumption and relative humidity or visibility (S3 Fig)

**Fig 4.**
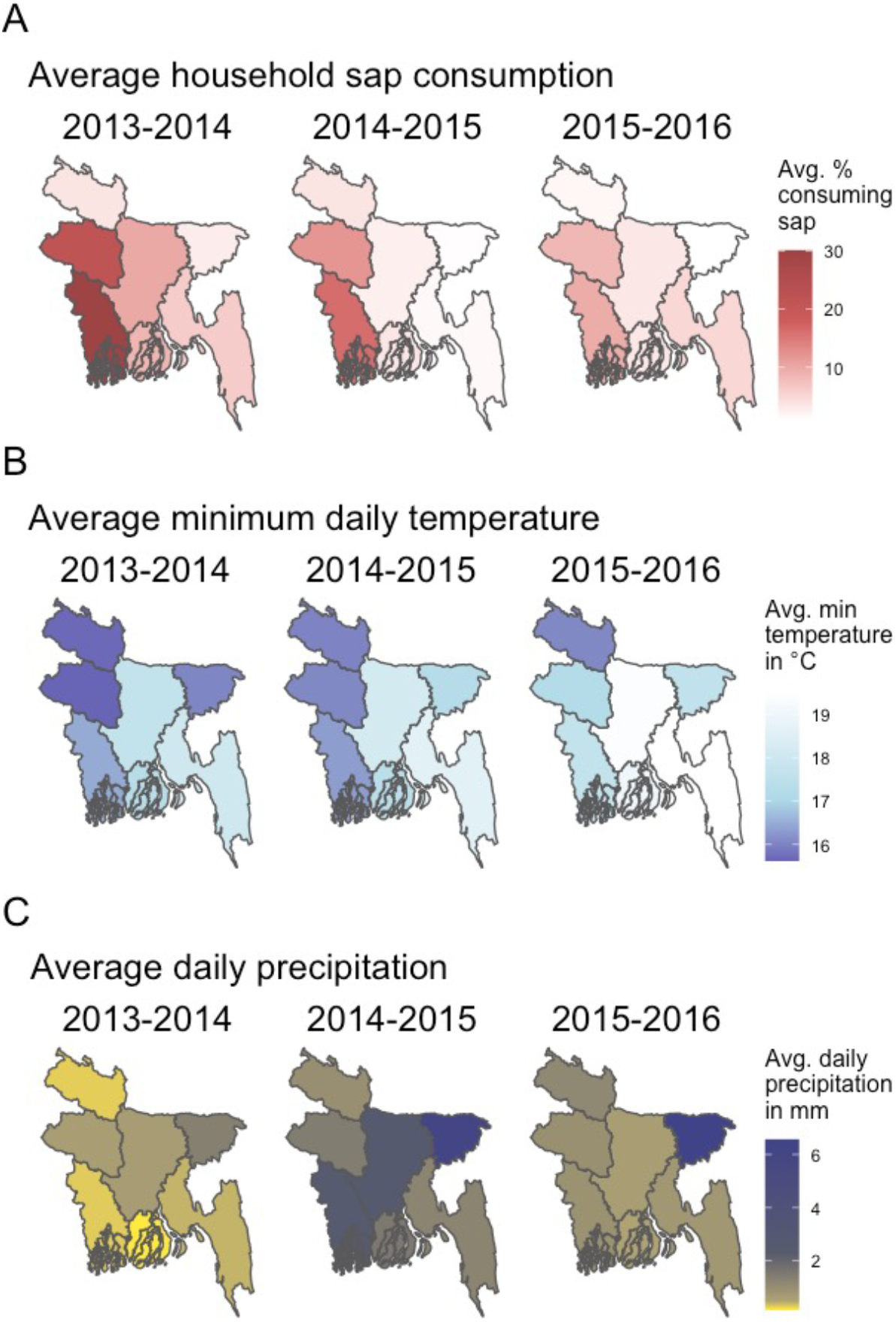
Sap consumption and weather by location and season. Relationship between date palm sap consumption and weather in Bangladesh, November-April, 2013-2016. (A) Average percentages of household sap consumption over sap seasons by division (B) Average of all daily minimum temperatures in °C over sap seasons by division (C) Average of all daily values of cumulative precipitation in millimeters over sap seasons by division.

We found that moving averages of minimum temperature contributed significantly to explaining sap consumption (**χ**^2^ (1, n =5,027) = 7.74, *p* < 0.01), though categorical monthly averages did not (**χ**^2^= 1.31, *p* = 0.252). A similar observation was made for cumulative precipitation, though both the moving (**χ**^2^ = 5.15, *p* < 0.05) and categorical (**χ**^2^ = 14.69, *p* < 0.001) averages improved model fit. Further, moving averages of cumulative precipitation contributed to model fit after accounting for the contributions of minimum temperature (**χ**^2^ = 8.00, *p* < 0.01), suggesting an independent association of each weather condition with sap consumption after accounting for the contributions of month, location, and sap season. Visibility or relative humidity was not found to be associated with household date palm sap consumption (S1 Table).

Comparing ROC curves for season and weather variables (S4 Fig), we found that there was no significant difference between the AUC when using only season (0.790) or only weather (0.786) as classifiers alongside month and location (Z = 0.562, *p* = 0.574, 95% CI: -0.011-0.0200). Examining the covariates of season and weather in model selection using AICc, the categorical variable of season provided better fit than moving averages of minimum temperature and precipitation, but the best fit was observed when including both season and weather (S2 Table and S4 Fig).

## Discussion

A study of human sap consumption during 2013-2016 in Bangladesh reported that consumption decreased across the study period and proposed multiple possible reasons for this decrease, including regional public health interventions advocating against sap consumption that were ongoing during the study period [15]. However, previous evidence highlighting the impacts of decreased temperature and precipitation on Nipah virus spillover [8,9, 14] and evidence suggesting both empiric and anecdotal support for the impacts of weather on date palm sap production [12, 13] support a proposed causal relationship between weather variation and sap consumption. We observed that household date palm sap consumption increased during colder, drier winters, and that relationship was largely generalizable across all months of the sap season and each administrative division in Bangladesh. Further, colder temperatures and less precipitation were associated with greater sap consumption even after accounting for sap season. Thus, we concur with the observation in Shanta et al. [15] that the reported decreases in sap consumption might be better explained by weather patterns during 2013-2016 rather than previous interventions. These findings indicate that weather-driven variability in sap consumption may be one of the major contributors to year-to-year variation in Nipah virus spillover.

Though identifying the association between winter weather patterns and sap consumption is useful in understanding interannual variability in Nipah virus spillover, the causal mechanisms of the association are still poorly understood. The effects of temperature and precipitation on the quantity or quality of sap produced by date palms remains unclear. It is possible that humans consume more sap during colder winters because the trees produce more sap during colder days, resulting in more opportunities for human consumption. Qualitative evidence from observers, historical records, and interviews with sap harvesters suggest that sap quality and volume may be highest on the coldest nights of the winter sap season [1, 12, 13]. Dilution of the sap from rain may visually impact the quality of the sap, reducing the likelihood that an individual would purchase it for consumption. Future studies into the effects of weather on date palm sap production quantity and quality could clarify the mechanisms that underlie the association between weather and date palm sap consumption. Additionally, as we do not have data on attitudes towards date palm sap consumption during the study period and thus cannot rule out the impact of education sap consumption interventions, future studies should take into account the motivating factors in the purchase of date palm sap beyond the quality of the sap itself.

Despite these unknowns, however, synthesizing previous evidence and identifying gaps in knowledge to confidently identify the behavioral and ecological processes between bat hosts, date palm trees, weather conditions, and human populations that drive Nipah seasonality exemplifies the benefit of an interdisciplinary approach to zoonotic disease. Utilizing approaches from ecology, virology, epidemiology, and anthropology, we can provide a uniquely broad view of a seasonal zoonotic disease system. Despite the challenge of understanding the numerous factors underlying zoonotic disease spillover, this approach may serve as a model for investigating the underlying interactions that determine seasonal variability in spillover events for other zoonotic pathogens.

## Data Availability

All data produced in the present study are available upon reasonable request to the authors.

https://www.ncei.noaa.gov/past-weather/

## References

1. Kamaluddin M, Nath TK, Jashimuddin M. Indigenous practice of Khejur palm (Phoenix sylvestris) husbandry in rural Bangladesh. Journal of Tropical Forest Science 1998;10:357–66.

2. Luby SP, Rahman M, Hossain M, Blum LS, Husain MM, Gurley ES, et al. Foodborne Transmission of Nipah Virus, Bangladesh. Emerg Infect Dis. 2006;12(12):1888–1894. doi: 10.3201/eid1212.060732.

3. Rahman MA, Hossain MJ, Sultana S, Homaira N, Khan SU, Rahman M, et al. Date Palm Sap Linked to Nipah Virus Outbreak in Bangladesh, 2008. Vector-Borne and Zoonotic Diseases 2012;12:65–72. doi: 10.1089/vbz.2011.0656.

4. Hegde ST, Sazzad HMS, Hossain MJ, Alam M-U, Kenah E, Daszak P, et al. Investigating Rare Risk Factors for Nipah Virus in Bangladesh: 2001–2012. EcoHealth 2016;13:720–8. doi: 10.1007/s10393-016-1166-0.

5. Luby SP. The pandemic potential of Nipah virus. Antiviral Research. 2013 Oct 1;100(1):38–43.

6. Nikolay B, Salje H, Hossain MJ, Khan AKMD, Sazzad HMS, Rahman M, et al. Transmission of Nipah Virus — 14 Years of Investigations in Bangladesh. New England Journal of Medicine 2019;380:1804–14. doi: 10.1056/NEJMoa1805376.

7. Khan, SUM, Hossain J, Gurley ES, Nahar N, Sultana R, Luby SP. Use of Infrared Camera to Understand Bats’ Access to Date Palm Sap: Implications for Preventing Nipah Virus Transmission. EcoHealth 2010;7:517–25. doi: 10.1007/s10393-010-0366-2.

8. McKee CD, Islam A, Luby SP, Salje H, Hudson PJ, Plowright RK, et al. The Ecology of Nipah Virus in Bangladesh: A Nexus of Land-Use Change and Opportunistic Feeding Behavior in Bats. Viruses 2021;13:169. doi: 10.3390/v13020169.

9. Cortes MC, Cauchemez S, Lefrancq N, Luby SP, Jahangir Hossain M, Sazzad HMS, et al. Characterization of the Spatial and Temporal Distribution of Nipah Virus Spillover Events in Bangladesh, 2007–2013. J Infect Dis 2018;217:1390–4. doi: 10.1093/infdis/jiy015.

10. Fogarty R, Halpin K, Hyatt AD, Daszak P, Mungall BA. Henipavirus susceptibility to environmental variables. Virus Res 2008;132:140–4. doi: 10.1016/j.virusres.2007.11.010.

11. Islam A, McKee C, Ghosh PK, Abedin J, Epstein JH, Daszak P, et al. Seasonality of Date Palm Sap Feeding Behavior by Bats in Bangladesh. EcoHealth 2021;18:359–71. doi: 10.1007/s10393-021-01561-9.

12. Nahar N, Sultana R, Gurley ES, Hossain MJ, Luby SP. Date Palm Sap Collection: Exploring Opportunities to Prevent Nipah Transmission. EcoHealth 2010;7:196–203. doi: 10.1007/s10393-010-0320-3.

13. Annett HE, Amin BM, Lele GK. The date sugar industry in Bengal : an investigation into its chemistry and agriculture. Calcutta: Published for the Imperial Dept. of Agriculture in India by Thacker, Spink & Co; 1913. doi: 10.5962/bhl.title.27998.

14. Latinne A, Morand S. Climate Anomalies and Spillover of Bat-Borne Viral Diseases in the Asia–Pacific Region and the Arabian Peninsula. Viruses 2022;14:1100. doi: 10.3390/v14051100.

15. Shanta IS, Luby SP, Hossain K, Heffelfinger JD, Kilpatrick AM, Haider N, Rahman T, Chakma S, Ahmed SSU, Sharker Y, Pulliam JRC, Kennedy ED, Gurley ES. Human Exposure to Bats, Rodents and Monkeys in Bangladesh. Ecohealth. 2023 Mar;20(1):53–64. doi: 10.1007/s10393-023-01628-9.

16. R Core Team (2021). R: A language and environment for statistical computing. R Foundation for Statistical Computing, Vienna, Austria. https://www.R-project.org/.

