## Supplemental Materials for "Identifying weather patterns affecting household date palm sap consumption in Bangladesh, 2013-2016"

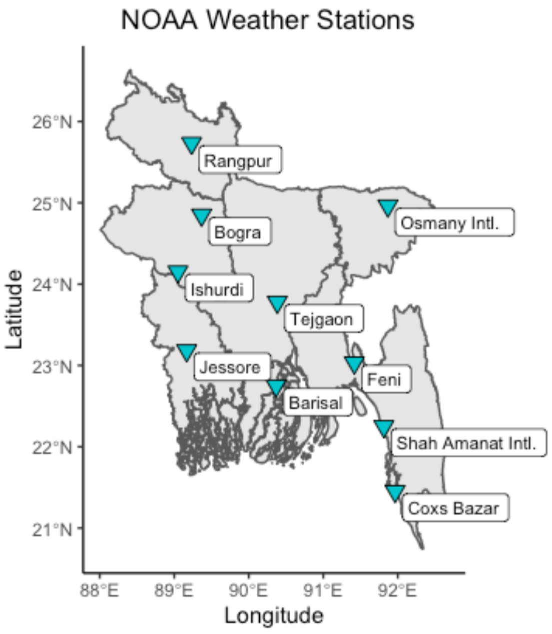


**S1 Fig**. Coverage of Bangladesh by NOAA weather stations.

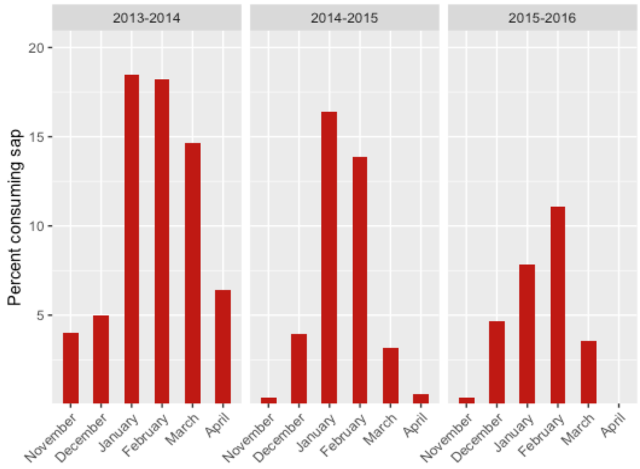

**S2 Fig.** Average percent household sap consumption in all locations by month over the study period.

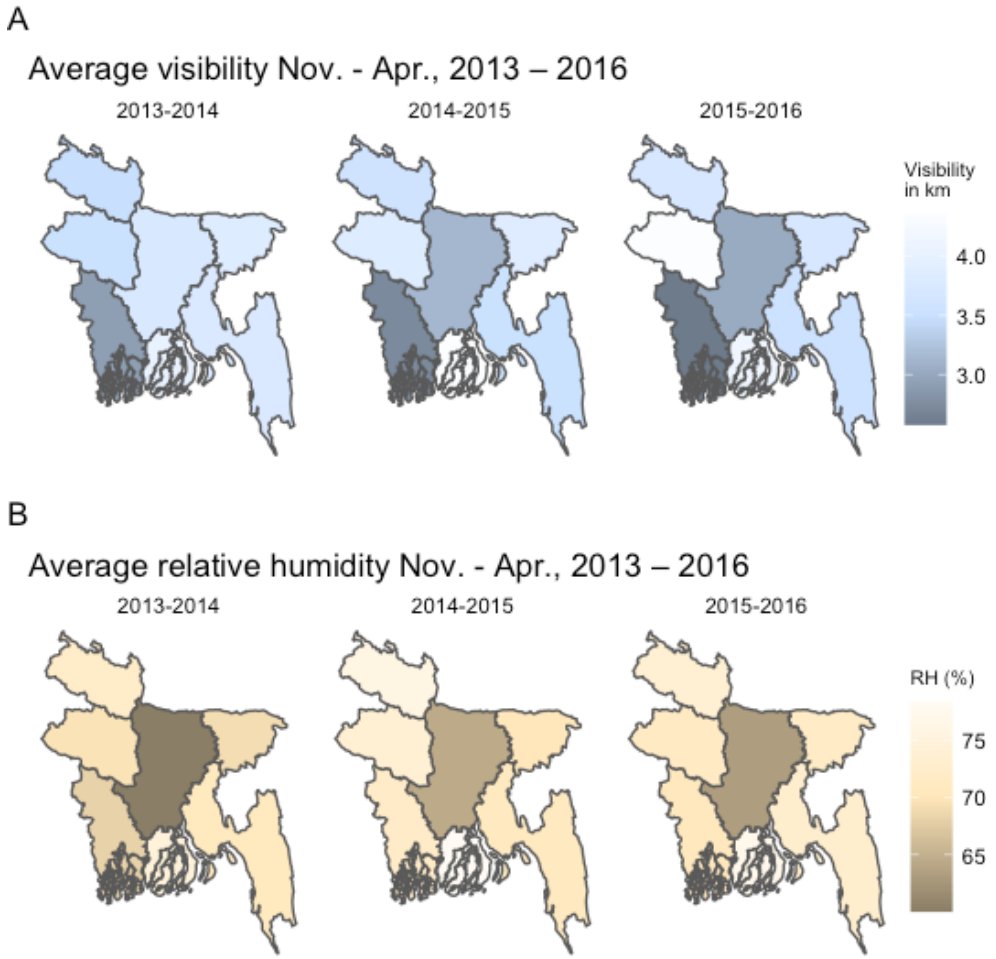


**S3 Fig. Visibility and relative humidity by location and season.** (A) Average of all daily values of visibility in kilometers over sap seasons by division (B) Average of all daily percentages of relative humidity over sap seasons by division.

**S1 Table**. $\chi^2$ Likelihood ratio tests to determine contribution of extended covariates to specified null models for household date palm sap consumption in the past month in responses given November- April 2013-2016 in Bangladesh.

| **Null** | **Extended** | **DF** | **LR $\chi^2$** |
|---|---|---|---|
| month + division | **+ season** | 1 | 56.36 *** |
| month + division + season | **+ cat_mintemp** | 1 | 1.31 |
| month + division + season | **+ SMA_mintemp** | 1 | 7.74 ** |
| month + division + season | **+ cat_precip** | 1 | 14.69 *** |
| month + division + season | **+ SMA_precip** | 1 | 5.15 * |
| month + division + season + SMA_mintemp | **+ cat_precip** | 1 | 13.75 *** |
| month + division + season + SMA_mintemp | **+ SMA_precip** | 1 | 8.00 ** |
| month + division + season + cat_precip | **+ SMA_mintemp** | 1 | 6.80 ** |
| month + division + season + SMA_precip | **+ SMA_mintemp** | 1 | 10.59 ** |
| month + division + season | **+ cat_visibility** | 1 | 0.23 |
| month + division + season | **+ SMA_visibility** | 1 | 2.86 |
| month + division + season | **+ cat_relativehumidity** | 1 | 1.13 |
| month + division + season | **+ SMA_relativehumidity** | 1 | 2.35 |

*= significant at $p < 0.05$
** = significant at $p < 0.01$
*** = significant at $p < 0.001$

cat: categorical monthly average of daily values for the weather variable within the division
SMA: simple moving average of daily values for the weather variable in the division within the 30 days preceding the response

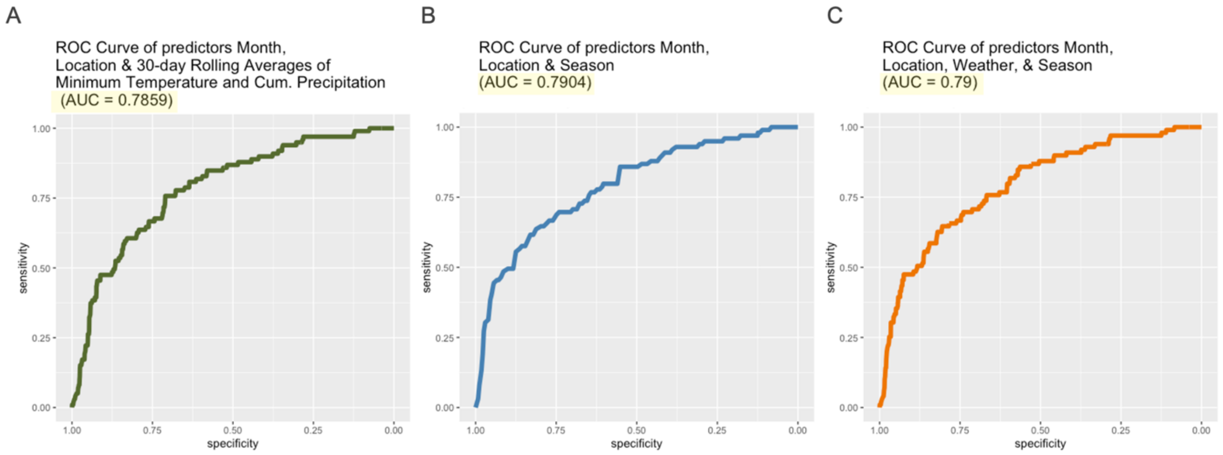


**S4 Fig. Receiver operating curves of specified models of weather, season, and sap consumption.** (A) Receiver operating curve of logistic regression model of month, location (administrative division), and 30-day rolling averages of minimum temperature and cumulative precipitation on date palm sap consumption, (B) Receiver operating curve of logistic regression model of month, location (administrative division), and sap season on date palm sap consumption, (C) Receiver operating curve of logistic regression model of month, location (administrative division), 30-day rolling averages of minimum temperature and cumulative precipitation, and sap season on date palm sap consumption.

**S2 Table.** Comparison of candidate logistic models of specified variables on date palm sap consumption using AICc.

| **Covariates** | **K** | **AICc** | **Δ AICc** | **Log-Likelihood** |
|---|---|---|---|---|
| month + division + season + SMA_mintemp + SMA_precip | 16 | **2119.19** | - | -1043.54 |
| month + division + season | 14 | **2130.91** | 11.71 | -1051.41 |
| month + division + SMA_mintemp + SMA_precip | 14 | **2133.98** | 14.79 | -1052.95 |
| month + division | 12 | **2183.25** | 64.46 | -1079.59 |

SMA: simple moving average of daily values for the weather variable in the division within the 30 days preceding the response
